# How feasible is it to mobilize $31 billion a year for pandemic preparedness and response? An economic growth modelling analysis

**DOI:** 10.1101/2023.10.29.23297740

**Authors:** Minahil Shahid, Marco Schäferhoff, Garrett Brown, Gavin Yamey

**Affiliations:** Center for Policy Impact in Global Health; Global Health Institute, Duke University; Open Consultants; University of Leeds

**Keywords:** pandemic preparedness and response, COVID-19, low- and middle-income countries, health financing

## Abstract

**Background:** Covid-19 has reinforced the strong health and economic case for investing in pandemic preparedness and response (PPR). The World Bank and World Health Organization (WHO) propose that low- and middle-income governments and donor countries should invest $31.1 billion each year for PPR. We analyse, based on the projected economic growth of countries between 2022 and 2027, how likely it is that low- and middle-income country governments and donors can mobilize the estimated funding.

**Methods:** We modelled trends in economic growth to project domestic health spending by low- and middle-income governments and official development assistance (ODA) by donors for years 2022 to 2027. We modelled two scenarios for countries and donors – a constant and an optimistic scenario. Under the constant scenario we assume that countries and donors continue to dedicate the same proportion of their health spending and ODA as a share of gross domestic product (GDP) and gross national income (GNI), respectively, as they did during baseline (the latest year for which data are available). In the optimistic scenario, we assume a yearly increase of 2.5% in health spending as a share of GDP for countries and ODA as a share of GNI for donors.

**Findings:** Our analysis shows that low-income countries would need to invest on average 37%, lower-middle income countries 9%, and upper-middle income countries 1%, of their total health spending on PPR each year under the constant scenario to meet the World Bank WHO targets. Donors would need to allocate on average 8% of their total ODA across all sectors to PPR each year to meet their target.

**Conclusions:** The World Bank WHO targets for PPR will not be met unless low- and middle-income governments and donors spend a much higher share of their funding on PPR. Even under optimistic growth scenarios, low-income and lower-middle income countries will require increased support from global health donors. The donor target cannot be met using the yearly increase in ODA under any scenario. If the country and donor targets are not met, the highest-impact health security measures need to be prioritized for funding. Alternative sources of PPR financing could include global taxation (e.g., on financial transactions, carbon, or airline flights), cancelling debt, and addressing illicit financial flows. There is also a need for continued work on estimating current PPR costs and funding requirements in order to arrive at more enduring and reliable estimates.

## Introduction

The covid-19 pandemic has had devastating health consequences, causing mass death, disability (e.g., from Long Covid), and orphanhood. The International Monetary Fund estimates that the economic losses caused by covid-19 will be close to US$ 13.8 trillion from 2020 to 2024 (1). Even before the pandemic, major gaps had been identified in the global health security architecture (2). Covid-19 has reinforced the strong health and economic case for investing in pandemic preparedness and response (PPR) (3). Such investments can help prevent, detect, and contain disease outbreaks, thereby reducing the broader social and economic costs of a pandemic (3,4).

How much would it cost to establish a global PPR system that is fully fit-for-purpose? Despite ongoing dialogue, there is currently no consistently applied approach to calculating global PPR resource requirements. (5) Previous estimates have ranged from US$ 1.6 billion to US$ 43 billion per year, depending on the costing methodology used, preparedness activities considered, and countries included in the analysis. (6)

The World Bank and the World Health Organization (WHO) recently provided a new estimate of the annual PPR financing needs in a report conducted for the G20 Joint Finance and Health Task Force. (4) The international community is now coalescing around these new figures. The World Bank and WHO estimate that low- and middle-income country governments and donors need to invest US$ 31.1 billion annually in PPR, of which US$ 26.4 billion needs to be invested at the country level and US$ 4.7 billion at the international level. (4,7) The report also acknowledges that low- and lower-middle income countries are unlikely to meet their national PPR financing requirements, estimating that there is an overall annual funding gap of US$ 10.5 billion at global and country levels. (4)

A critical question to answer is: assuming the figure is correct, how feasible is it to achieve this annual “price tag” of US$ 31.1 billion? We therefore set out to address this question by analysing, based on the economic growth that the International Monetary Fund projects for years 2022 to 2027 (8), how likely it is that low- and middle-income countries and donors will mobilize the estimated funding for PPR.

Considering various scenarios, we addressed two questions. First, how realistic is it for low- and middle-income governments to reach the annual country level PPR finance target of US$ 26.4 billion from growth in domestic health spending? Second, how feasible is it for donors to support low- and middle-income countries to reach this US$ 26.4 billion target, while at the same time financing global (international-level) PPR needs? In addition, we challenge the US$ 10.5 billion annual funding gap identified by the World Bank/WHO, suggesting that it is based on poor assumptions, and that the funding gap is actually closer to US$ 15.5 billion. Box 1 summarizes how we defined low- and middle-income countries and donors.

### Box 1

**Definition of low- and middle-income countries and donors**

We conducted separate analyses for low- and middle-income countries and donors:

- **Low- and middle-income countries**. We used the World Bank’s classification of countries by income group (9) to identify and conduct assessment for: low-income countries (LICs), lower-middle-income countries (LMICs), and upper-middle-income countries (UMICs). In total, we included 115 countries in our analysis (Appendix 1). Seventeen countries were excluded due to lack of data.
- **Donor countries**. We identified donor countries using the Organisation for Economic Co-operation and Development (OECD) Disbursements and Commitments of Official and Private Flows Statistics Database (10,11). Our analysis included a total of 43 donor countries—of which 29 are members of the OECD Development Assistance Committee (DAC) (12) and 14 are non-DAC donors (Appendix 2). Five donor nations (Azerbaijan, Chinese Taipei, Kazakhstan, Kuwait, and Liechtenstein) were excluded due to lack of data.

## Methods

Our study did not obtain an ethics approval as it does not involve human subjects and only uses publicly available national and international financial data and published secondary resources.

### Low- and Middle-Income Countries’ Analysis

Using low- and middle-income country gross domestic product (GDP) data from the World Bank (13) for the year 2021, we projected the GDP for each country from 2022 to 2027 using the annual percentage change in GDP data from the International Monetary Fund’s (IMF) World Economic Outlook (WEO) database (8). We used real GDP to account for the effects of inflation across all analyses. After calculating the projected economic growth (in constant 2020 US$) for low- and middle-income countries, we used the ‘Domestic General Government Health Expenditure (GGHE-D) (also referred to as ‘domestic health spending’) as a percent of GDP’ data from the WHO Global Health Expenditure database (14), and multiplied these values with the projected GDP data from the WEO dataset to get the projected domestic health spending by low- and middle-income countries for the years 2022 to 2027. For the projected years, we calculated two different scenarios:

1. <u>Constant Scenario:</u> We assumed that the share of government health expenditures out of GDP remained constant (i.e. we assumed that the latest available ‘GGHE-D as a percent of GDP’ data, which are currently for the year 2020, apply to all years from 2022 to 2027). In other words, in this constant scenario, low- and middle-income countries continue to spend the same percentage of their GDP on domestic health spending between the years 2022 to 2027 as they did in 2020.
2. <u>Scale-up Scenario:</u> We increased the ‘GGHE-D as a percent of GDP’ ratio by 2.5 percent each year up until 2027. This is a more optimistic scenario, one in which low- and middle-income countries recognize the need to marginally increase the percentage of their GDP spent on health year on year.

For both scenarios, we projected the trend in GGHE-D for each income group and calculated what share of GGHE-D would be required to meet the annual US$ 26.4 billion PPR target (Table 1 shows the cost breakdown by income group). In addition, we examined the yearly increment in domestic health spending under the two scenarios to estimate what proportion of the increment would be needed to meet the PPR target. Our analysis focuses on the anticipated growth in GGHE-D, and it does not examine any redistribution of domestic health spending from other priority areas such as infectious disease control or maternal and child health. We used GDP data from the World Bank (13) and calculations were performed in 2020 US$.

**Table 1:** Estimated national-level annual PPR target by income group.

| Income Group | Cost (Billion US\$) | Cost (%) |
| --- | --- | --- |
| LIC Target | 2.7 | 10.2% |
| LMIC Target | 13.5 | 51.1% |
| UMIC Target | 10.2 | 38.6% |
| **Total** | **26.4** | **100.0%** |

**Table 2:**
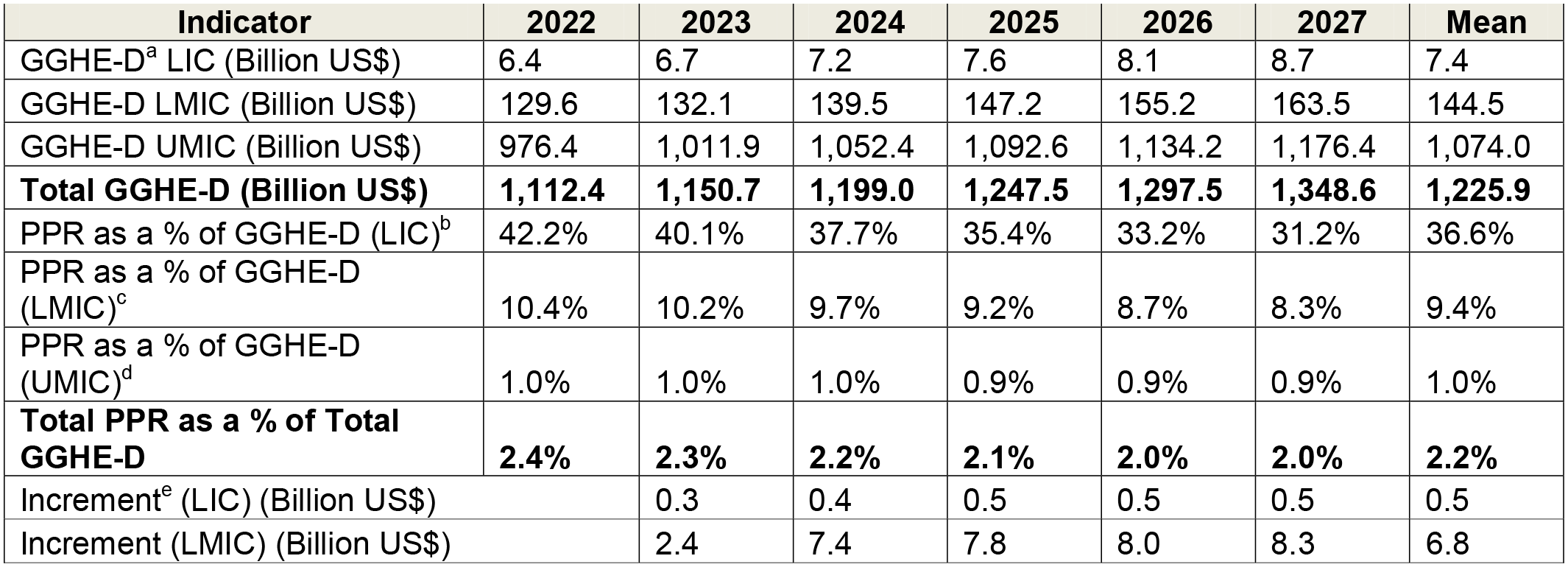
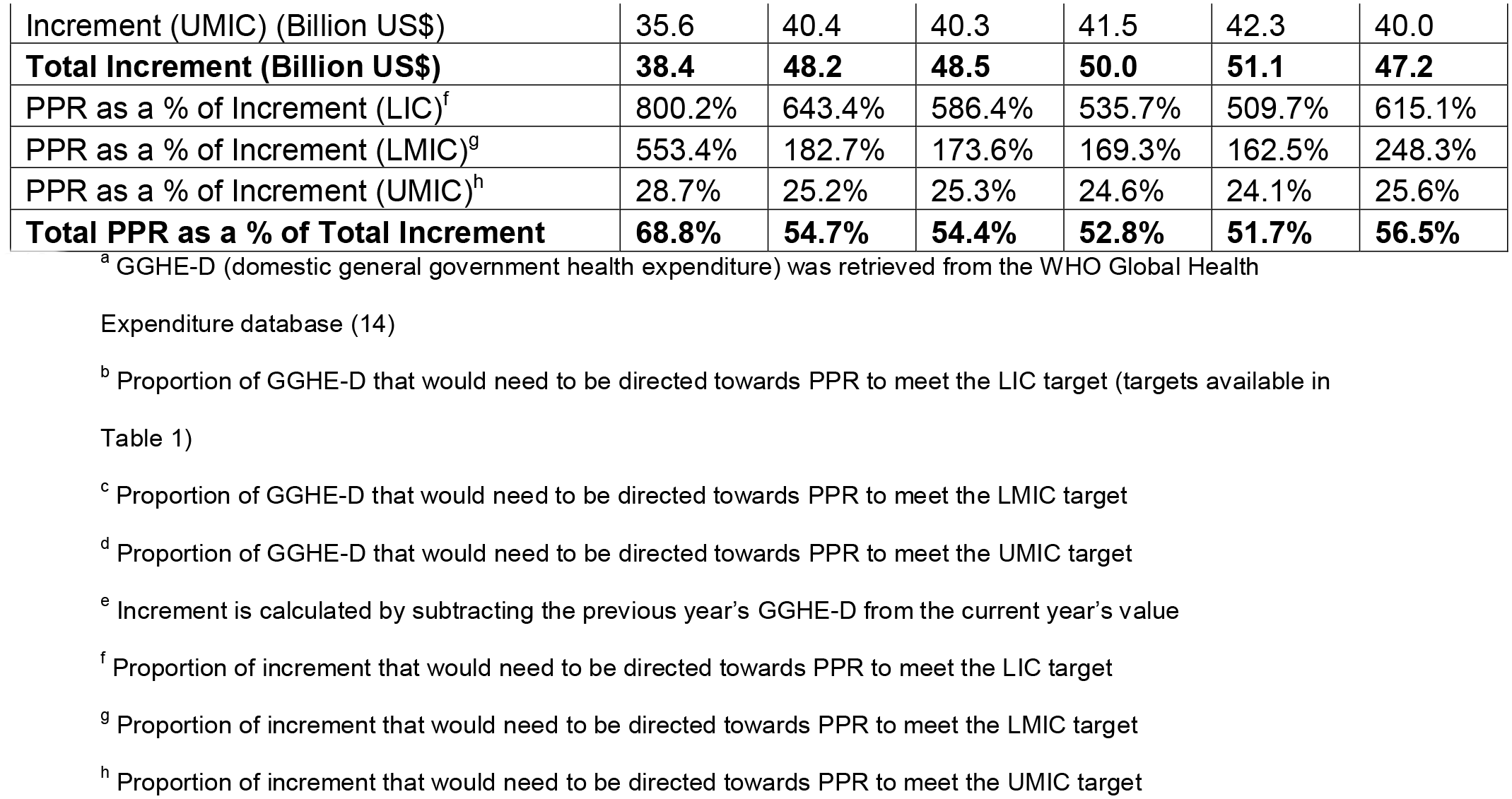
Projected growth in domestic health spending by low- and middle-income country governments under the constant scenario.

### Donor Analysis

We estimated the increase or decrease in official development assistance (ODA) that donor countries are projected to give between the years 2022 and 2027 using projected changes to their economic growth (i.e., changes in their gross national income [GNI]). We used GNI rather than GDP due to limitations in data availability; there is a relatively small difference in values between the two indicators (i.e., using GNI or GDP would give similar results).

We used the annual percentage change in GDP data for years 2022-2027 from the IMF’s WEO database. We multiplied this percentage change with the Gross National Income (GNI) data for donor countries for the year 2021 from the OECD DAC1 (10) database to calculate the projected economic growth for DAC and non-DAC donors for the years 2022 to 2027.

The OECD DAC1 database provides historical data on disbursements and commitments of official and private flows from members of the Development Assistance Committee (DAC), multilateral organisations and other donors.

We then used the ‘Official Development Assistance (ODA) as a percent of GNI’ data from the OECD DAC1 database to calculate the projected ODA flows by donors for the years 2022 to 2027. We created two scenarios (similar to the country-level analyses) described below:

1. <u>Constant Scenario:</u> Keeping the ODA/GNI ratio constant, we assumed that donors continue to give the same percent of their GNI to ODA between 2022 to 2027 as they did in 2021; in this scenario, any increase or decrease in ODA is solely driven by changes in donors’ GNI data;
2. <u>Scale-Up Scenario:</u> We assume that there is a yearly 2.5 percent increase in the 2021 ODA to GNI ratio, compounding to a total increase of 15 percent by 2027.

To capture different scenarios, we varied the proportion of GNI that donor nations might give to ODA between 2022-27 using the 2021 ODA/GNI ratio as the baseline (the latest year for which ODA/GNI ratio data were available). A pessimistic scenario with a decreasing ODA-to-GNI ratio can potentially stem from budgetary pressures and ODA cuts by certain donor countries. In contrast, an optimistic view would suggest that, for example, given the economic and social losses caused by covid-19, donors find value in investing a greater proportion of their GNI towards ODA, leading to an overall increase in ODA availability. In line with our country-level GGHE-D analysis, we projected ODA growth from 2022-2027 under two scenarios – a constant and a scale-up scenario.

In our analysis, we assume that donors would cover the entire global-level PPR investment of US$ 4.7 billion, and also provide 100% of the annual PPR funding for LICs (US$ 2.7 billion per year) and 60% for LMICs (US$ 8.1 billion per year). The assumptions on the share of funding covered by donors reflects those in the World Bank/WHO costing study. In total, we thus assume donors would need to provide an annual amount of US$ 15.5 billion.

## Results

### Low- and Middle-Income Countries Analysis Results

Our analysis shows that LICs, under the constant scenario, would have to invest 42.2% of their total annual GGHE-D to reach the PPR target in 2022, while in 2027 they would still need to spend 31.2% of their GGHE-D on PPR (Table 3). Under the scale-up scenario, LICs would have to invest 26.9% of their GGHE-D in 2027 to meet the PPR target (Table 4). The increment in funding under both constant and scale-up scenarios is insufficient for reaching the PPR target for LICs.

**Table 3:**
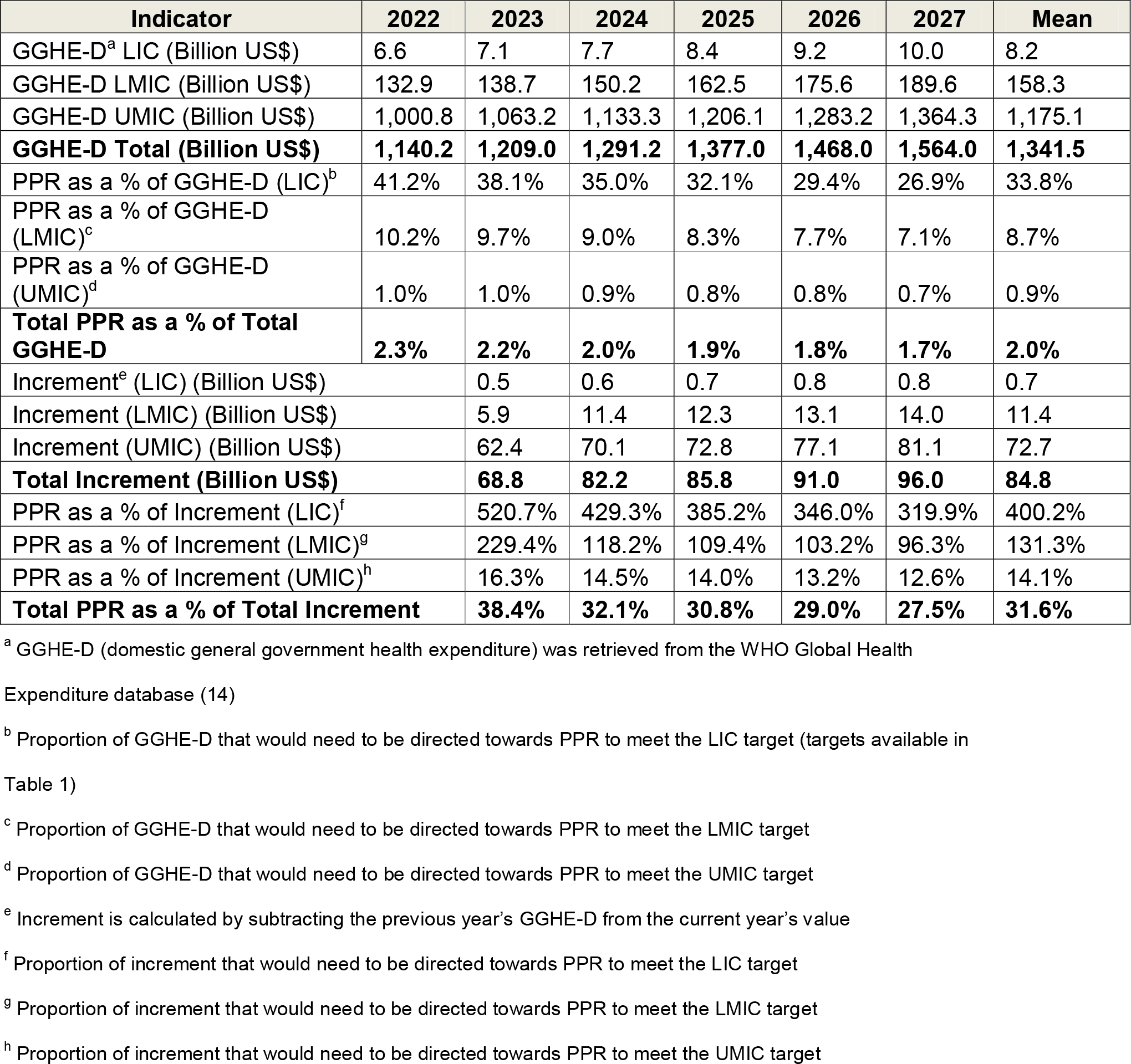
Projected growth in domestic health spending by low- and middle-income country governments under the scale-up scenario.

**Table 4:** Projected growth in official development assistance by donors under the constant and scale-up scenarios.

|  | 2022 | 2023 | 2024 | 2025 | 2026 | 2027 | Mean |
| --- | --- | --- | --- | --- | --- | --- | --- |
| **Constant Scenario** |  |  |  |  |  |  |  |
| ODA (Billion US\$) | 186.2 | 190.5 | 193.7 | 197.0 | 200.2 | 203.3 | 195.1 |
| PPR as a % of ODA<sup>a</sup> | 8.3% | 8.1% | 8.0% | 7.9% | 7.7% | 7.6% | 7.9% |
| Increment (Billion US\$) | 7.4 | 4.2 | 3.2 | 3.3 | 3.2 | 3.1 | 4.1 |
| Increment as a % of PPR<sup>b</sup> | 48.0% | 27.4% | 20.9% | 21.1% | 20.5% | 19.9% | 26.3% |
| **Scale-Up Scenario** |  |  |  |  |  |  |  |
| ODA (Billion US\$) | 190.9 | 200.1 | 208.6 | 217.5 | 226.5 | 235.7 | 213.2 |
| PPR as a % of ODA<sup>a</sup> | 8.1% | 7.7% | 7.4% | 7.1% | 6.8% | 6.6% | 7.3% |
| Increment (Billion US\$) | 12.1 | 9.2 | 8.5 | 8.8 | 9.0 | 9.2 | 9.5 |
| Increment as a % of PPR<sup>b</sup> | 78.0% | 59.6% | 54.8% | 57.0% | 58.2% | 59.6% | 61.2% |

<sup>a</sup> Proportion of ODA that would need to be directed towards PPR to meet the US\$ 15.5 billion target each year
<sup>b</sup> Proportion of annual US\$ 15.5 billion target that would be met if the entire increment was directed at PPR

For LMICs, the percentage of GGHE-D needed to meet their PPR target drops from 10.4% in 2022 to 8.3% in 2027 under the constant scenario and to 7.1% in the scale-up scenario. The increment in projected funding is also insufficient for LMICs to meet their PPR costs.

For UMICs, the annual PPR costs account for 1.0% of their total annual GGHE-D in the constant scenario, and drop to 0.7% in 2027 under the scale-up scenario. UMICs, unlike LICs and LMICs, would be able to cover PPR costs using a substantial share of their incremental funding – 25.6% on average under the constant scenario and 14.1% on average under the scale-up scenario.

A limitation of our analysis stems from the composition of the GGHE-D indicator in the WHO Global Health Expenditure database. The indicator does not account for capital expenditures while calculating domestic health spending (15), which leads to an overall underestimation in projected GGHE-D. We therefore conducted a sensitivity analysis (see Appendix 3), and included capital expenditures (reported separately in the WHO database) in our analysis.

### Donor Analysis Results

Our analysis shows that the US$ 15.5 billion annual PPR target for donor government spending would not fully be met under any scenario, even if the entire yearly increase in ODA is used for PPR. Even under the scale-up scenario of a 15% linear increase in the ODA/GNI ratio, and even if the entire yearly increase in ODA over the six years is directed to PPR, it would only cover an annualised average of 61% of the US$ 15.5 billion annual PPR requirement (see Table 4), and 90% if the target was brought down to US$ 10.5 billion.

## Discussion

Our analysis for the constant scenarios shows that LICs would need to invest on average 37%, LMICs 9%, and UMICs 1%, of their total health spending on PPR each year to meet the World Bank WHO targets. Donors would need to allocate on average 8% of their total ODA across all sectors to PPR each year to meet their target.

Based on these projections, we believe it is not feasible for low- and lower-middle-income governments to reach their annual PPR funding targets from domestic spending alone. Even under the optimistic scenario, LICs would still have to allocate, on average, 34% of their total GGHE-D between 2022 and 2027 to PPR, making the target untenable. The largest PPR costs relate to LMICs – a diverse group of countries with variable abilities to pay their own PPR needs. Our analysis shows that LMICs would need to spend on average 7-10% of their GGHE-D on PPR between 2022 and 2027.

In terms of the increment, under the constant scenario, the increment resulting from economic growth is insufficient for LMICs to meet their PPR requirements (the increment for LMICs, on average from 2022 to 2027, is US$ 6.8 billion, while the requirement is US$ 13.5 billion per year). One potential implication is that LMICs would need to reduce their health spending on other priorities to meet the target.

In addition, many LMICs will lose support from the Global Fund, Gavi, the Vaccine Alliance, and other donors in the coming years and will need to increase their domestic spending on priorities such as HIV, tuberculosis, malaria, and vaccination programs.(16) Their likely economic growth will allow them to increase their health spending, but given competing priorities it is unrealistic to suggest that more than the entire growth in projected health funding will be directed towards PPR because that would imply a reallocation from other health areas. UMICs are able to finance their own PPR target, and only need to dedicate on average 1% of their GGHE-D, or 14-26% of their increment, to PPR.

It is clear that donors would need to support LICs and LMICs. Given the constraints of these country groups to self-finance their PPR needs, it is important to be realistic and transparent about the amount of additional donor funding required. The World Bank/WHO costing study argues that the annual priority need at country-level is US$ 26.4 billion, of which US$ 16.2 billion or 61.3% would fall on LICs and LMICs, while the remainder is for UMICs, who, according to the World Bank, can cover those costs themselves. Because the World Bank/WHO study assumes that donors already cover 100% and 60% of the LIC and LMIC costs respectively, the annual funding gap at country level is reduced from $26.4 billion to $7.0 billion.

However, we challenge the assumption that donors already provide such a significant amount of funding for PPR to LICs and LMICs in non-pandemic times. Although health ODA significantly increased in 2020 and 2021 (from US$ 22.2 billion in 2019 to US$ 29.2 billion in 2020 and US$ 34.0 billion in 2021), much of this increase can be attributed to the covid-19 response, especially donor funding for covid-19 vaccines. (17) In previous years, donors invested very little in PPR. (18) In addition, there is evidence that existing ODA and national level resources for health have shifted to COVID-19 and PPR activities, signalling a reallocation of scarce resources which can threaten existing Universal Health Coverage (UHC) vulnerabilities. (19) As a result, the assumptions of the World Bank/WHO study about existing donor funding appear to be unrealistic.

We argue that the annual donor funding needed amounts to US$15.5 billion – US$ 4.7 billion for global and regional PPR, US$ 2.7 billion per year for LICs and US$ 8.1 billion per year for LMICs. Calculating the annual gap thus requires more realistic data and assumptions on donor spending for PPR during non-pandemic times as well as a recognition that the annual funding gap is much higher than US$ 10.5 billion.

How feasible is it to mobilize US$15.5 billion in donor funding for PPR every year through 2027? Donors would need to allocate 7-8% of their total ODA – across all sectors - to PPR between 2022 and 2027. In terms of the increment in total ODA, on average, the increment could cover 26% and 61% of the PPR requirement under the constant and scale-up scenarios, respectively. In other words, even if the entire increase in ODA increment is spent on PPR, which is unrealistic in itself, donors would not be able to meet this funding without sufficient increases in the percentage of ODA allocated to PPR via new funding. Ideally, this would not merely be a redistribution from other ODA commitments.

How might the global health community respond to these projections? One approach is to simply accept these projections and design plans for how to efficiently spend whatever financing does get mobilized by identifying and prioritising the highest value for money and PPR impact measures. Another, and potentially complementary approach, would be for donors to increase their funding beyond the 2.5% ODA/GNI ratio as a PPR investment strategy against the type of public health and economic risk experienced during covid-19. Here, there are arguments to be made from a benefit/cost perspective that could make ODA/GNI increases more palatable to donors and their constituents.

Moreover, aside from reprioritizing domestic health spending and ODA towards PPR, alternative sources of financing outside the usual health related sources should be explored (e.g., from national security or defence budgets) (20). For example, there is growing interest in levying a global tax on financial transactions, carbon, or airline flights to help fund PPR (21). Debt cancellation must also be on the table. Public debt in LMICs increased from 58% to 65% of GDP from 2019 to 2021. (22) The cost of borrowing for low-income countries has also increased compared to pre-pandemic levels and is projected to continue increasing as global interest rates rise. (22) If the G20 and financial institutions had cancelled all external debt payments due in 2020 and 2021 by the 76 poorest countries, it would have liberated US$ 300 billion (23).

Addressing illicit financial flows (IFFs) and global tax abuses that continue to trickle wealth from low- and middle-income countries into higher-income nations could also help redirect resources from illicit channels into more productive ones such as investing in PPR. Countries with high IFFs are reported to spend 25% less on health compared to countries with low IFFs. (24) Eastern and Southern Africa lost US$ 7.6 billion in tax revenue in 2017, equivalent to 1.6% of the region’s GDP, due to only two sources of IFFs (base erosion and profit shifting to tax havens). (25) Addressing IFFs is important, since countries with high IFFs are reported to spend 25% less on health compared to countries with low IFFs. (24) Measures to tackle IFFs can strengthen LMIC financial systems and also free resources for public health purposes.

Reducing the cost of PPR itself is also desirable and would require strong measures including reducing constraints on intellectual property (IP) to allow equitable global access to safe and affordable medical countermeasures (MCMs). During covid-19, pharmaceutical companies partnered with certain high-income countries to hinder IP waiver negotiations, stalling progress towards equitable access to covid-19 vaccines. (26,27) If new global PPR initiatives such as the Pandemic Fund and the Pandemic Treaty are to be successful, they must facilitate technology transfer, IP waivers, and support local manufacturing of medical countermeasures to help lower PPR resource requirements.

## Data Availability

We used publicly available datasets for our analysis. These include
International Monetary Fund World Economic Outlook database, Organisation for Economic Co-operation and Development (OECD) Disbursements and Commitments of Official and Private Flows Statistics Database (DAC1, CRS), World Health Organization Global Health Expenditure Database, World Bank World Development Indicators

