## Appendix 1 for "How feasible is it to mobilize $31 billion a year for pandemic preparedness and response? An economic growth modelling analysis"

Countries included in the analysis of LMIC government domestic health spending:

| Burundi | Comoros | Honduras | Mongolia |
| --- | --- | --- | --- |
| Algeria | Costa Rica | India | Montenegro |
| Angola | Côte d'Ivoire | Indonesia | Morocco |
| Argentina | Democratic Republic of the Congo | Iraq | Mozambique |
| Armenia | Djibouti | Jamaica | Myanmar |
| Azerbaijan | Dominica | Jordan | Namibia |
| Bangladesh | Dominican Republic | Kazakhstan | Nepal |
| Belarus | Ecuador | Kenya | Nicaragua |
| Belize | Egypt | Kyrgyz Republic | Niger |
| Benin | El Salvador | Lao P.D.R. | Nigeria |
| Bolivia | Equatorial Guinea | Lesotho | North Macedonia |
| Bosnia and Herzegovina | Eswatini | Liberia | Pakistan |
| Botswana | Ethiopia | Madagascar | Papua New Guinea |
| Brazil | Fiji | Malawi | Paraguay |
| Bulgaria | Gabon | Malaysia | Peru |
| Burkina Faso | Georgia | Maldives | Philippines |
| Cabo Verde | Ghana | Mali | Russia |
| Cambodia | Grenada | Marshall Islands | Rwanda |
| Cameroon | Guatemala | Mauritania | Samoa |
| Central African Republic | Guinea | Mauritius | São Tomé and Príncipe |
| Chad | Guinea-Bissau | Mexico | Senegal |
| China | Guyana | Micronesia | Serbia |
| Colombia | Haiti | Moldova | Sierra Leone |

LMICs not included in the analysis:

| Cook Islands | Islamic Republic of Iran |
| --- | --- |
| Niue | Lebanon |
| Palau | Congo |
| Tonga | Afghanistan |
| Turkmenistan | Yemen |
| Albania | South Sudan |
| Cuba | Eritrea |
| Kiribati | Venezuela |
| Bhutan |  |
