## Appendix 2 for "How feasible is it to mobilize $31 billion a year for pandemic preparedness and response? An economic growth modelling analysis"

Countries included in the analysis of donor government spending on ODA:

*DAC donors included:*

| Australia | Korea |
| --- | --- |
| Austria | Luxembourg |
| Belgium | Netherlands |
| Canada | New Zealand |
| Czech Republic | Norway |
| Denmark | Poland |
| Finland | Portugal |
| France | Slovak Republic |
| Germany | Slovenia |
| Greece | Spain |
| Hungary | Sweden |
| Iceland | Switzerland |
| Ireland | United Kingdom |
| Italy | United States |
| Japan |  |

*Non-DAC donors included:*

Bulgaria

Croatia

Cyprus

Estonia

Israel

Latvia

Lithuania

Malta

Qatar

Romania

Saudi Arabia

Thailand

Turkey

United Arab Emirates

Countries not included in the donor analysis:

*Non-DAC donors not included:*

Azerbaijan

Chinese Taipei

Kazakhstan

Kuwait

Liechtenstein
