## Appendix 3 for "How feasible is it to mobilize $31 billion a year for pandemic preparedness and response? An economic growth modelling analysis"

Sensitivity Analysis (including Capital Expenditures data)

Table 5: Projected Growth in Domestic Health Spending by Low- and Middle-Income Country Governments under the Constant Scenario (including Capital Expenditures)

| **Indicator** | **2022** | **2023** | **2024** | **2025** | **2026** | **2027** | **Mean** |
| --- | --- | --- | --- | --- | --- | --- | --- |
| Capital Exp.^a^ LIC (Billion US$) | 1.0 | 1.1 | 1.2 | 1.2 | 1.3 | 1.4 | 1.2 |
| Capital Exp. LMIC (Billion US$) | 20.4 | 20.8 | 22.0 | 23.4 | 24.7 | 26.1 | 22.9 |
| Capital Exp. UMIC (Billion US$) | 128.2 | 134.3 | 140.7 | 147.3 | 154.1 | 161.1 | 144.3 |
| **Total Capital Exp. (Billion US$)** | **149.7** | **156.1** | **163.9** | **171.9** | **180.2** | **188.7** | **168.4** |
| PPR as a % of GGHE-D + Cap. Exp (LIC) ^b^ | 36.3% | 34.4% | 32.4% | 30.4% | 28.5% | 26.7% | 31.5% |
| PPR as a % of GGHE-D + Cap. Exp. (LMIC)^c^ | 9.0 % | 8.8% | 8.4% | 7.9% | 7.5% | 7.1% | 8.1% |
| PPR as a % of GGHE-D + Cap. Exp. (UMIC)^d^ | 0.9% | 0.9% | 0.9% | 0.8% | 0.8% | 0.8% | 0.8% |
| **Total PPR as a % of Total GGHE-D + Cap. Exp.** | **2.1%** | **2.0%** | **1.9%** | **1.9%** | **1.8%** | **1.7%** | **1.9%** |
| Increment Cap. Exp.^e^ (LIC) (Billion US$) | | 0.1 | 0.1 | 0.1 | 0.1 | 0.1 | 0.1 |
| Increment Cap. Exp (LMIC) (Billion US$) | | 0.3 | 1.3 | 1.3 | 1.4 | 1.4 | 1.1 |
| Increment Cap. Exp (UMIC) (Billion US$) | | 6.0 | 6.5 | 6.6 | 6.8 | 7.0 | 6.6 |
| **Total Increment Cap. Exp. (Billion US$)** | | **6.4** | **7.8** | **8.0** | **8.3** | **8.5** | **7.8** |
| PPR as a % of Increment (GGHED + Cap. Exp.) (LIC)^f^ | | 686.1% | 549.1% | 502.4% | 450.5% | 428.4% | 523.3% |
| PPR as a % of Increment (GGHED + Cap. Exp.) (LMIC)^g^ | | 492.2% | 155.8% | 148.0% | 144.5% | 139.1% | 215.9% |
| PPR as a % of Increment (GGHED + Cap. Exp.) (UMIC)^h^ | | 24.5% | 21.8% | 21.8% | 21.1% | 20.7% | 22.0% |
| **Total PPR as a % of Total Increment (GGHED + Cap. Exp.)** | | **59.0%** | **47.1%** | **46.7%** | **45.3%** | **44.3%** | **48.5%** |

^a^ Cap. Exp. (capital expenditure) data was retrieved from the WHO Global Health Expenditure database (14)

^b^ Proportion of GGHE-D and capital expenditures that would need to be directed towards PPR to meet the LIC target (targets available in Table 1)

^c^ Proportion of GGHE-D and capital expenditures that would need to be directed towards PPR to meet the LMIC target

^d^ Proportion of GGHE-D and capital expenditures that would need to be directed towards PPR to meet the UMIC target

^e^ Increment is calculated by subtracting the previous year’s capital expenditures from the current year’s value

^f^ Proportion of capital expenditure increment and GGHE-D increment that would need to be directed towards PPR to meet the LIC target

^g^ Proportion of capital expenditure increment and GGHE-D increment that would need to be directed towards PPR to meet the LMIC target

^h^ Proportion of capital expenditure increment and GGHE-D increment that would need to be directed towards PPR to meet the UMIC target

Table 6: Projected Growth in Domestic Health Spending by Low- and Middle-Income Country Governments under the Scale-Up Scenario (including Capital Expenditures)

| **Indicator** | **2022** | **2023** | | **2024** | **2025** | **2026** | **2027** | **Mean** |
| --- | --- | --- | --- | --- | --- | --- | --- | --- |
| Capital Exp. ^a^ LIC (Billion US$) | 1.1 | 1.2 | | 1.3 | 1.4 | 1.5 | 1.7 | 1.3 |
| Capital Exp. LMIC (Billion US$) | 21.0 | 21.8 | | 23.7 | 25.8 | 28.0 | 30.3 | 25.1 |
| Capital Exp. UMIC (Billion US$) | 131.4 | 141.1 | | 151.5 | 162.6 | 174.4 | 186.8 | 158.0 |
| **Total Capital Exp.** **(Billion US$)** | **153.5** | **164.0** | | **176.5** | **189.8** | **203.9** | **218.8** | **184.4** |
| PPR as a % of GGHE-D + Cap. Exp (LIC)^b^ | 35.4% | 32.8% | | 30.1% | 27.6% | 25.2% | 23.1% | 29.0% |
| PPR as a % of GGHE-D + Cap. Exp. (LMIC)^c^ | 8.8% | 8.4% | | 7.8% | 7.2% | 6.6% | 6.1% | 7.5% |
| PPR as a % of GGHE-D + Cap. Exp. (UMIC)^d^ | 0.9% | 0.8% | | 0.8% | 0.7% | 0.7% | 0.7% | 0.8% |
| **Total PPR as a % of Total GGHE-D + Cap. Exp** | **2.0%** | **1.9%** | | **1.8%** | **1.7%** | **1.6%** | **1.5%** | **1.8%** |
| Increment Cap. Exp. (LIC)^e^ (Billion US$) | | | 0.1 | 0.1 | 0.1 | 0.1 | 0.2 | 0.1 |
| Increment Cap. Exp. (LMIC) (Billion US$) | | | 0.8 | 1.9 | 2.1 | 2.2 | 2.3 | 1.9 |
| Increment Cap. Exp. (UMIC) (Billion US$) | | | 9.6 | 10.5 | 11.1 | 11.8 | 12.4 | 11.1 |
| **Total Increment Cap. Exp. (Billion US$)** | | | **10.6** | **12.5** | **13.2** | **14.1** | **14.9** | **13.1** |
| PPR as a % of Increment (GGHED + Cap. Exp.) (LIC)^f^ | | | 446.8% | 367.1% | 330.3% | 292.6% | 270.3% | 341.4% |
| PPR as a % of Increment (GGHED + Cap. Exp.) (LMIC)^g^ | | | 200.7% | 101.2% | 93.7% | 88.4% | 82.6% | 113.3% |
| PPR as a % of Increment (GGHED + Cap. Exp.) (UMIC)^h^ | | | 14.2% | 12.7% | 12.2% | 11.5% | 10.9% | 12.3% |
| **Total PPR as a % of Total Increment (GGHED + Cap. Exp.)** | | | **33.3%** | **27.9%** | **26.6%** | **25.1%** | **23.8%** | **27.3%** |

^a^ Cap. Exp. (capital expenditure) data was retrieved from the WHO Global Health Expenditure database (14)

^b^ Proportion of GGHE-D and capital expenditures that would need to be directed towards PPR to meet the LIC target (targets available in Table 1)

^c^ Proportion of GGHE-D and capital expenditures that would need to be directed towards PPR to meet the LMIC target

^d^ Proportion of GGHE-D and capital expenditures that would need to be directed towards PPR to meet the UMIC target

^e^ Increment is calculated by subtracting the previous year’s capital expenditures from the current year’s value

^f^ Proportion of capital expenditure increment and GGHE-D increment that would need to be directed towards PPR to meet the LIC target

^g^ Proportion of capital expenditure increment and GGHE-D increment that would need to be directed towards PPR to meet the LMIC target

^h^ Proportion of capital expenditure increment and GGHE-D increment that would need to be directed towards PPR to meet the UMIC target
